# Mapping the environmental co-benefits of reducing low-value care: a scoping review and bibliometric analysis

**DOI:** 10.1101/2024.04.23.24306217

**Authors:** Gillian Parker, Sarah Hunter, Karen Born, Fiona A. Miller

## Abstract

**Background:** Reducing low-value care (LVC) and improving healthcare’s climate readiness are critical factors to improve the sustainability of health systems across the globe. Care practices that have been deemed low or no value, in effect, generate carbon emissions, waste and pollution without improving patient or population health. There is nascent, but growing, research and evaluation to inform practice change focused on the environmental co-benefits of reducing LVC. The objective of this study was to develop foundational knowledge of this field through a scoping review and bibliometric analysis.

**Methods:** We searched four databases, Medline, Embase, Scopus and CINAHL, each from inception to July 2023. We followed established scoping review and bibliometric analysis methodology to collect and analyze the data. Publication characteristics, healthcare and environmental sustainability focus (scoping review); authors, institutions, institution countries, and collaborations (bibliometric analysis) data were collected.

**Findings:** 145 publications met inclusion criteria and were published between 2013 – July 2023; with over 80% published since 2020. Empirical studies represented 21% while commentary, editorials or opinions represented 51% of publications. The majority focused on healthcare generally (27%), followed by laboratory testing (14%), and medications (14%). Empirical publications covered a broad range of environmental issues with general and practice-specific ‘Greenhouse gas (GHG) emissions’, ‘waste management’ and ‘resource use’ as most common topics. Reducing practice-specific ‘GHG emissions’ was the most common reported environmental outcome. The bibliometric analysis revealed numerous international collaboration networks of prolific authors producing work across healthcare practices and settings, studying numerous environmental sustainability issues.

**Conclusions:** This study reveals that research and evaluation to inform practice change on the environmental co-benefits of reducing LVC is growing internationally, across multiple healthcare and environmental areas. Results demonstrate a need and opportunity for the emerging community to clarify approaches and strengthen the evidence-base through further empirical work in the field.

## INTRODUCTION

Health systems are significant contributors to climate change and other ecological harms, and are increasingly recognizing their obligation to mitigate their environmental impacts in the short-term while moving toward health system sustainability in the long term [1–3]. The most significant environmental impacts of healthcare come not from healthcare facilities themselves, but rather from the products and services that healthcare organizations deliver and consume [2, 4]. In 2023 at the COP28 Summit, 120 countries committed to putting health at the centre of climate action and flagging the need for low carbon sustainable and climate resilient health systems [5]. Strategies identified in the literature as critical to ensuring an effective and efficient health system include reducing low-value care (LVC) [6, 7] and reducing the environmental impact of healthcare [8–10]. Moreover, awareness is developing among clinicians, health system leaders and decision-makers of the co-benefits that can be realized through interventions that integrate efforts to reduce the environmental impacts of care with efforts to reduce (LVC) [11–13].

Low-value care includes care practices (tests, treatments or procedures) that have been identified, using scientific evidence, to be unnecessary, ineffective or harmful in hospital, primary care, long-term care or public health contexts [14]. Common examples of LVC include antibiotics for viral infections and laboratory testing prior to low-risk surgeries [7]. Reducing LVC offers myriad benefits including: improving patient care and outcomes and freeing resources for expanded coverage [7, 15–17]. Patients can be harmed by LVC, whether during the test or procedure or through infections, reactions or by over-testing or over treatment [7, 17, 18]. By definition, LVC generates carbon emissions, waste, and pollution without improving patient or population health [15, 19]. In addition, LVC occurs most often in high-income country (HIC) settings—settings where access and resources are abundant [20] while the environmental impact of LVC, in the form of GHG emissions for example, disproportionally impacts marginalized groups and those in low- and middle-income country (LMIC) settings [12, 21].

Within the climate change literature, “co-benefits” include the positive environmental impacts that a policy or intervention aimed at one objective might have on other objectives, thereby increasing the total benefit for society [22]. For health systems, addressing the challenge of LVC [11, 13] has the potential to be a critically important strategy for securing environmental co-benefits at the frontline of care delivery and at organization- and system-levels. Approaches to reducing LVC that identify, address, measure and report on environmental co-benefits have the potential to produce “win-win” situations and to overcome silos to achieve objectives of improved health and reduced environmental impact. Co-benefits, within the climate and environmental sustainability literature, have been described as “happy accidents” that produce a benefit. There may be potential to deliberately optimize such benefits by understanding interdependent relationships, identifying synergies, and addressing potential barriers [23]. In early 2023 the National Institute for Health and Care Excellence (NICE)—the UK National Health Service organization that publishes health technology, clinical, health promotion and social care guidelines—acknowledged the environmental co-benefits of reducing LVC as a critical health system strategy: “NICE can influence both direct and indirect carbon emissions, and already does so through its guidance and advice products and work to reduce the use of low value care.” [24p4].

### Objectives & Research Questions

While recognition and activity in this area of research and practice change is flourishing, to our knowledge, no reviews of the literature on the environmental co-benefits of reducing LVC have been published. The objectives of this study were to identify and characterize a body of literature to build foundational knowledge and advance understanding of this field through a scoping review and bibliometric analysis.

The research questions that motivated this study were: (Scoping Review) *How has knowledge production in this field evolved over time? What are the key and emerging areas of focus in this field?* (Bibliometric Analysis) *What are the prolific collaborations in this field? Who/what are the prolific authors, institutions, countries and publication sources in this field?*

## METHODS

We selected a scoping review and bibliometric analysis as the ideal methods to conduct our inquiry. Scoping reviews, a literature synthesis-type, are most appropriate when examining emerging and/or broad topics with the aim of characterizing their features [25]. Bibliometric analysis involves descriptive, statistical analysis of aggregated bibliometric metadata associated with relevant publications to provide insights into key topics, contributors (authors, author institutions and institution countries)—and relationships between them—of a particular research area (field) [26–29]. We used the scoping review methodological framework proposed by Arksey and O’Malley [25] and later enhanced by Levac, Colquhoun, and O’Brien [30]. The Preferred Reporting Items for Systematic Reviews and Meta-Analysis Extension for Scoping Reviews (PRISMA-ScR) [31] also informed the conduct and reporting of the scoping review component (see Supplementary File 1). Currently, no reporting checklist exists to support the conduct and/or reporting of bibliometric analysis. Accordingly, we developed and completed a reporting checklist, specific to bibliometric analysis, based on our knowledge of and experiences with the methodology (see Supplementary File 2).

### Search Strategy

A preliminary review of the literature was conducted to explore publications focused on the environmental co-benefits of reducing LVC and to inform the development of the search strategy. Based on this, we developed a 44-search term strategy and conducted a comprehensive search of four database: Scopus, MEDLINE, and EMBASE, and CINAHL (see Supplementary File 3). This search was first run in January 2023, capturing publications from each databases’ inception to January 2023, and then re-run in July 2023 to update the search to July 1, 2023. Search terms were categorized into three groups:

1. *Low-value care* (overdiagnos* or “low value” or low-value or overtest* or over-test* or overtreat* or over-treat* or “choosing wisely” or “less is more” or “reducing waste” or overuse or “minimal benefit*” or de-implement* or deimplement* or deadopt* or de-adopt* or de-prescrib* or unnecessary or “over surveillance”) AND;
2. *Environmental sustainability* (“environmentally sustainable” or “environmental sustainability” or “carbon footprint*” or “carbon emission*” or “climate change” or “net zero” or “climate risk” or “low carbon” or carbon* or de-carbon* or “carbon performance” or “indirect carbon impact*” or “environmental impact” or “GHG emission*” or “environmental emission*” or “greenhouse gas emission*” or carbon-constrained or “climate crisis”) AND;
3. *Health* (health* or healthcare or medicine* or medica* or “commissioned care” or hospital* or laborator*).

### Inclusion Criteria

Included publications focused (to varying degrees) on environmental co-benefits of reducing LVC. This focus could be primarily on reducing environmental harms of healthcare, with reducing LVC as a strategy or primarily on reducing LVC with a recognition of the environmental co-benefits or (approximately) equally focused. These criteria were purposefully broad as this is a nascent field and no literature was available regarding the scope of the field or possible data collection parameters. Included publications were not restricted by geography or healthcare setting. All types of publications were included (e.g., empirical studies, reviews, commentaries, editorials, conference abstracts) to capture all relevant knowledge production and comprehensively map its breadth and scope. Due to resource constraints, only English-language publications were included.

### Exclusion Criteria

Publications were excluded if they focused on non-human subjects, such as animals or agriculture, or were focused on natural or applied sciences (e.g., chemistry, earth science, engineering).

### Publication Selection

Search results were imported into Covidence, a Cochrane technology platform (www.covidence.org), and duplicates were automatically removed. Title and abstract screening and full-text review of publications were conducted by three research team members (GP, SH, TB) who resolved discrepancies through discussion. Two researcher (GP and SH) screened all titles and abstracts independently. One research team member (GP) reviewed a random 10% sample of screened abstracts and resolved discrepancies. Full-text review was conducted by two researchers (GP and SH), discrepancies were discussed and resolved collaboratively. The reference lists of included publications were hand searched to identify additional, relevant publications.

## SCOPING REVIEW-SPECIFIC METHODS

### Data Collection

The scoping review data collection worksheet was designed iteratively; piloted with 10 included publications and revised accordingly. Data collected included publication characteristics (publication year, type of publication), healthcare focus and environmental sustainability focus.

### Data Analysis

Data were entered into an Excel-based spreadsheet to facilitate data analysis and reporting. Data analysis was conducted by two research team members. Descriptive statistics were used to summarize the data. The data were analyzed by three members of the research team (GP, SH and FM) with discrepancies resolved collaboratively. All members of the research team reviewed the final summary of results.

### LVC and/or Environmental Sustainability Focus

While all publications included the environmental co-benefits of reducing LVC, the degree and amount of focus varied significantly. For example, some publications focused primarily on reducing the environmental impacts of healthcare and listed reducing LVC as a strategy, while others focused on reducing LVC with a recognition of the environmental co-benefits, finally, a subset focused on both aspects equally, reporting both reducing LVC and environmental sustainability outcomes. To document this characteristic, we categorized the publications in three groups:

1. Publications (approximately) equally focused on environmental sustainability of healthcare AND reducing LVC;
2. Publications primarily focused on environmental sustainability of healthcare (and acknowledging the benefits of reducing LVC); and,
3. Publications primarily focused on reducing LVC (and acknowledging the environmental co-benefits of doing so).

### Healthcare Focus

Categorizing the healthcare focus of these publications was complex as publications crossed disciplines, medical practices and healthcare settings. An inductive process was used by one researcher (GP) to map the categories and sub-categories, then validated by two research team members (SH and FM). Our categorization demonstrates the breadth and scope of healthcare focus in this field, but is not definitive nor necessarily exhaustive. We developed categories across four areas of healthcare focus (with 15 sub-categories): ‘*Procedures’* (Laboratory testing, imaging, respiratory, anaesthesia, surgery, Intensive Care Unit (ICU)*), ‘System organization/design/evaluation’* (general healthcare, measurement/metrics)*, ‘Pharmaceuticals’* (antibiotics and other) and *‘Care Type/Setting’* (hospital, primary, mental health psychiatry, nursing, other).

### Environmental Focus

As this is an emerging field, we decided to only collect data related to environmental sustainability for the subset of empirical papers we categorized as (approximately) equally focused on environmental sustainability of healthcare and reducing LVC. Our rationale was that we wanted to understand which aspects of environmental sustainability were being studied empirically when both environmental sustainability and reducing LVC were studied as a common focus (rather than parenthetically attending to one or other outcome).

Categorizing the environmental sustainability focus was also complex as publications often used vague or differing definitions of relevant terms and focused on broad areas of environmental sustainability. An inductive process was used by one researcher (GP) to map the categories and sub-categories, then validated by two research team members (SH and FM). We developed categories across six areas of environmental sustainability (with 16 sub-categories): ‘*GHG emissions’* (resulting from both general healthcare and specific practices), *‘Pollution’* (air, land, water), *‘Resource use’* (use less single-use products, water, energy/electricity, natural /non-renewable/ raw resources), *‘Waste management’*(Recycling, compostability, re-use, re-process, produce less solid waste, produce less waste (general), *‘Supply chain & facility/service design’*(service design/ processes, facilities/building/ system, sustainable supply chain. sustainable procurement), *and ‘Environmental stewardship’* (Education (decision-makers, providers, patients, public) and support/ influence/ actions of staff, suppliers, etc.).

As work on extending the breadth and scope of environmental sustainability addressed in healthcare is growing in prominence, our goal was to collect data on both primary (outcomes) and secondary (evidence cited or recommendations made) information from these empirical studies. We developed two groupings for this information and, within each of the sub-categories, we collected data and classified it as either: (i) *evidence* or r*ecommendation*, if a study cited existing evidence or made a recommendation in an environmental sustainability category; or (ii) *reported outcomes*, if a study reported new study outcomes in an environmental sustainability category.

## BIBLIOMETRIC ANALYSIS-SPECIFIC METHODS

### Data Collection

Raw metadata were retrieved for all publications analyzed in the scoping review, primarily from Web of Science (WoS) (per bibliometric analysis guidance) [27] with additional data retrieved from Embase, Scopus, and Medline as necessary.

### Data Cleaning

The raw metadata were converted in preparation for data cleaning, analysis, and visualization using Biblioshiny [27]. Data cleaning ensured the quality of data related to author names, author institutions (affiliate/s the author (and their work) is associated with at the time of publishing), publication sources (see Supplementary File 4 and File 5). Data cleaning was facilitated using Microsoft Excel and OpenRefine (https://openrefine.org/) software applications.

### Data Analysis and Visualization

Two analytic techniques were employed: publication [28], and co-authorship analysis [26]. Microsoft Excel, OpenRefine, and Biblioshiny [27] software applications were used to facilitate data analysis and visualization. Microsoft Excel- and Biblioshiny-generated data visualizations were recreated, and additional layers of analysis were applied as necessary, using the vector-based graphics software, Adobe Illustrator (see Supplementary File 4).

## RESULTS

### Literature search

The database searches identified 2456 publications (after duplicates were removed) for which the titles and abstracts were screened for inclusion. Of these, 372 publications were selected for full-text screening and 145 publications were included in these analyses (see Supplementary File 6 and File 7.). See Figure 1. a flow diagram we developed, inspired by the PRISMA flow diagram, representing the processes of publication selection, data collection, pre-processing, analysis, synthesis, visualization, and reporting for both the scoping review and bibliometric analysis.

**Figure 1.**
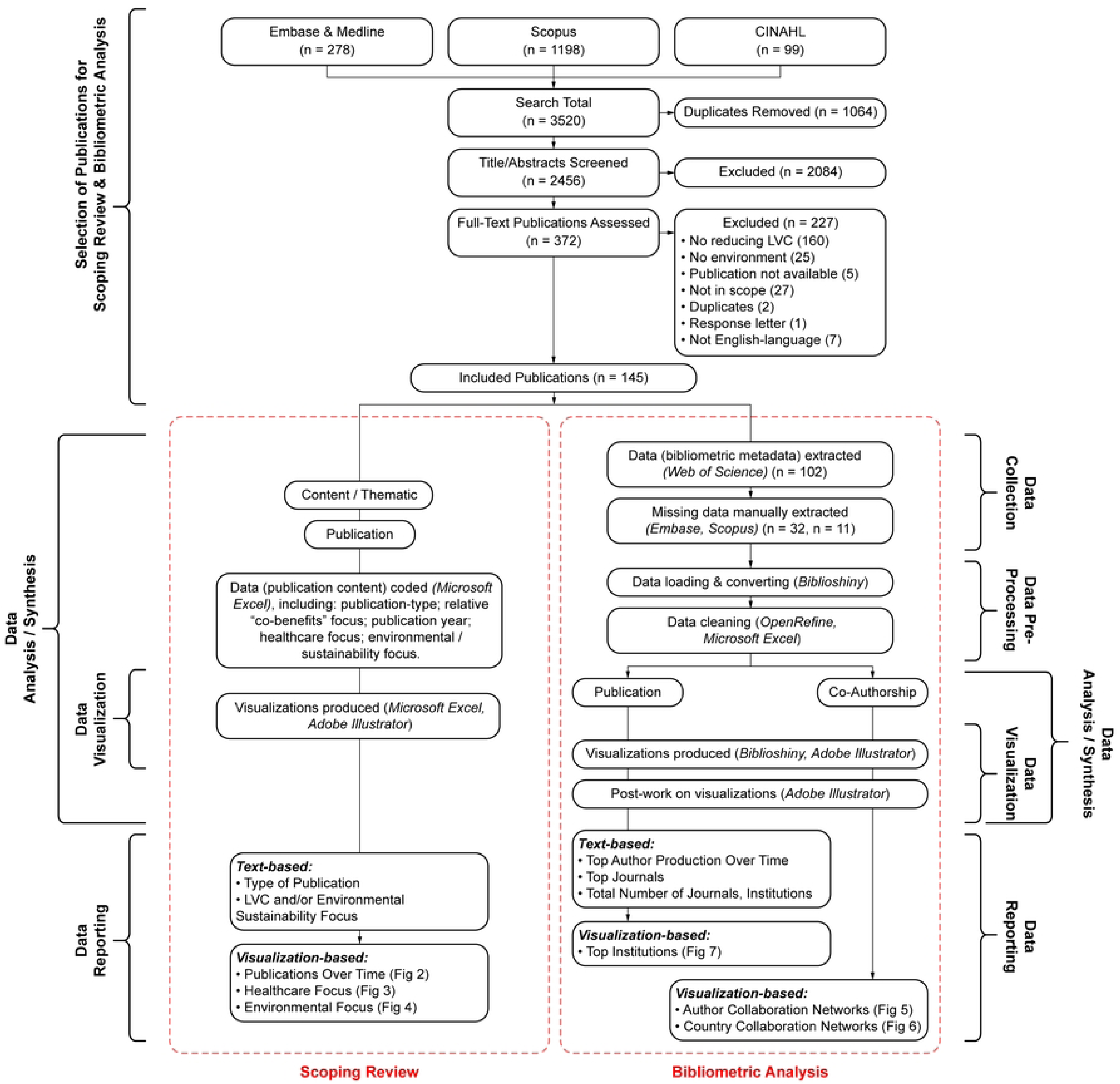
Flow diagram outlining the study design.

## SCOPING REVIEW-SPECIFIC RESULTS

### Publication Timeline

The first included paper was published in 2013. The majority of included publications (34%) were published within the first half of 2023, with only 12% of publications were produced before 2020 (Figure 2.).

**Figure 2.**
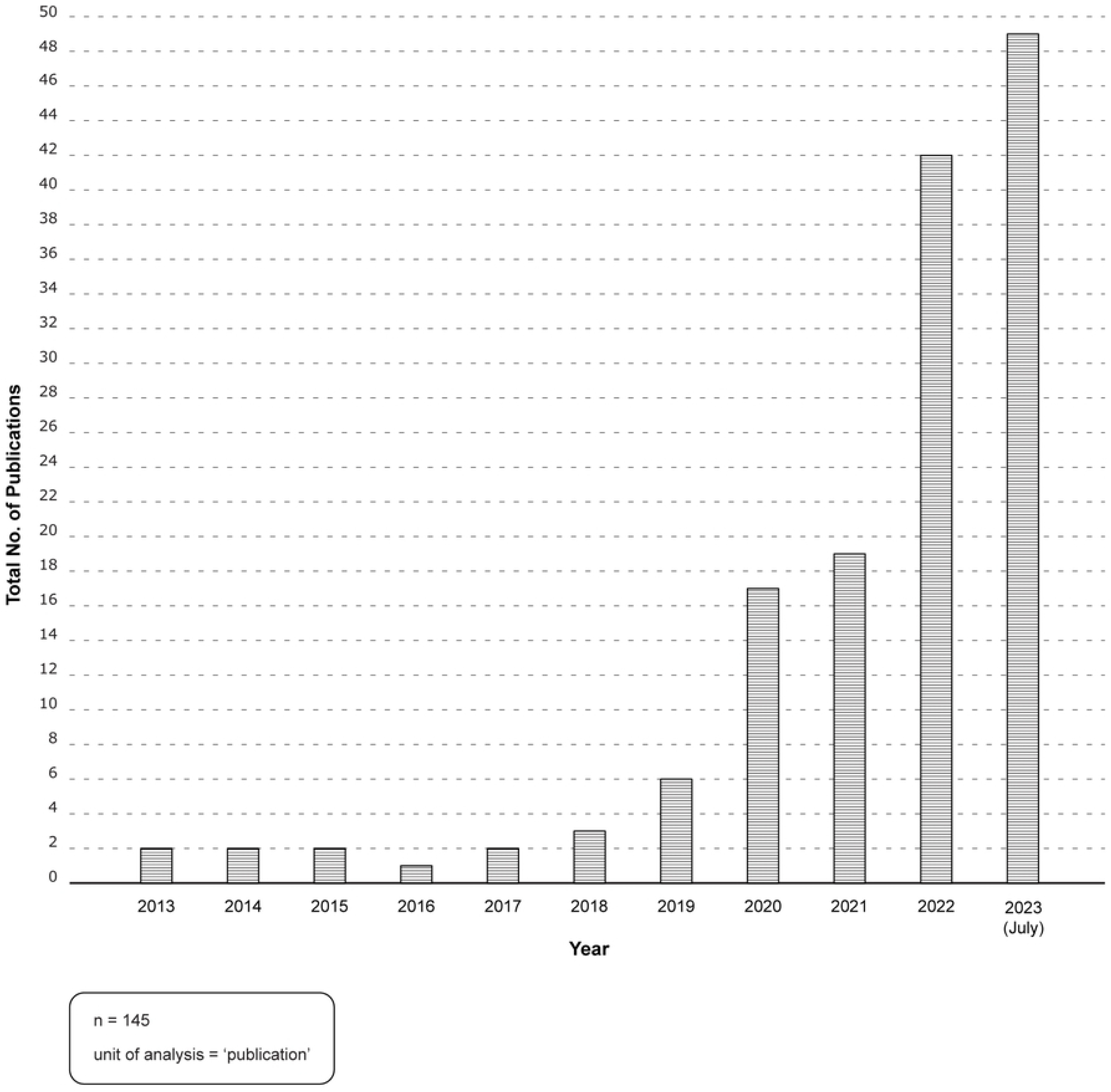
Publications over time. Source: Microsoft Excel: analysis; Adobe Illustrator: visualization.

### Type of Publication

The most prominent publication-type was commentaries/opinions/editorials/viewpoints (51%); followed by reviews (23%) and empirical studies (21%). The remaining 5% of publications included protocols, conference abstracts and position statements.

### LVC and/or Environmental Sustainability Focus

The majority of the publications were primarily focused on environmental sustainability of healthcare, with reducing LVC as a suggested strategy (52%); followed by publications (approximately) equally focused on both environmental sustainability and reducing LVC (34%); followed by publications primarily focused on reducing LVC with a recognition of the environmental co-benefits (14%).

### Healthcare Focus of Publications

Healthcare focus was recorded in four categories – ‘*Procedures’, ‘System organization/ design/ evaluation’, ‘Pharmaceuticals’ and ‘Care Type/Setting’* and 15 sub-categories (Figure 3.). The first category *‘Procedures’*, captured the majority of publications (42%) and included procedures related to the following subcategories: laboratory testing, imaging, respiratory, anaesthesia, surgery and ICU. The second category ‘*System organization/ design/ evaluation’* captured publications (30%) focused on the health system or healthcare generally or publications related to metrics or measurement. The third category *‘Pharmaceuticals’* (14%) was split into antibiotics and other pharmaceuticals to demonstrate the amount of work published specific to antibiotics. The fourth category *‘Care Type/Setting’* captured publications (14%) focused on care within a particular setting rather than a specific procedure or pharmaceutical. This category included primary care, hospital, mental health/psychiatry and nursing focused publications.

**Figure 3.**
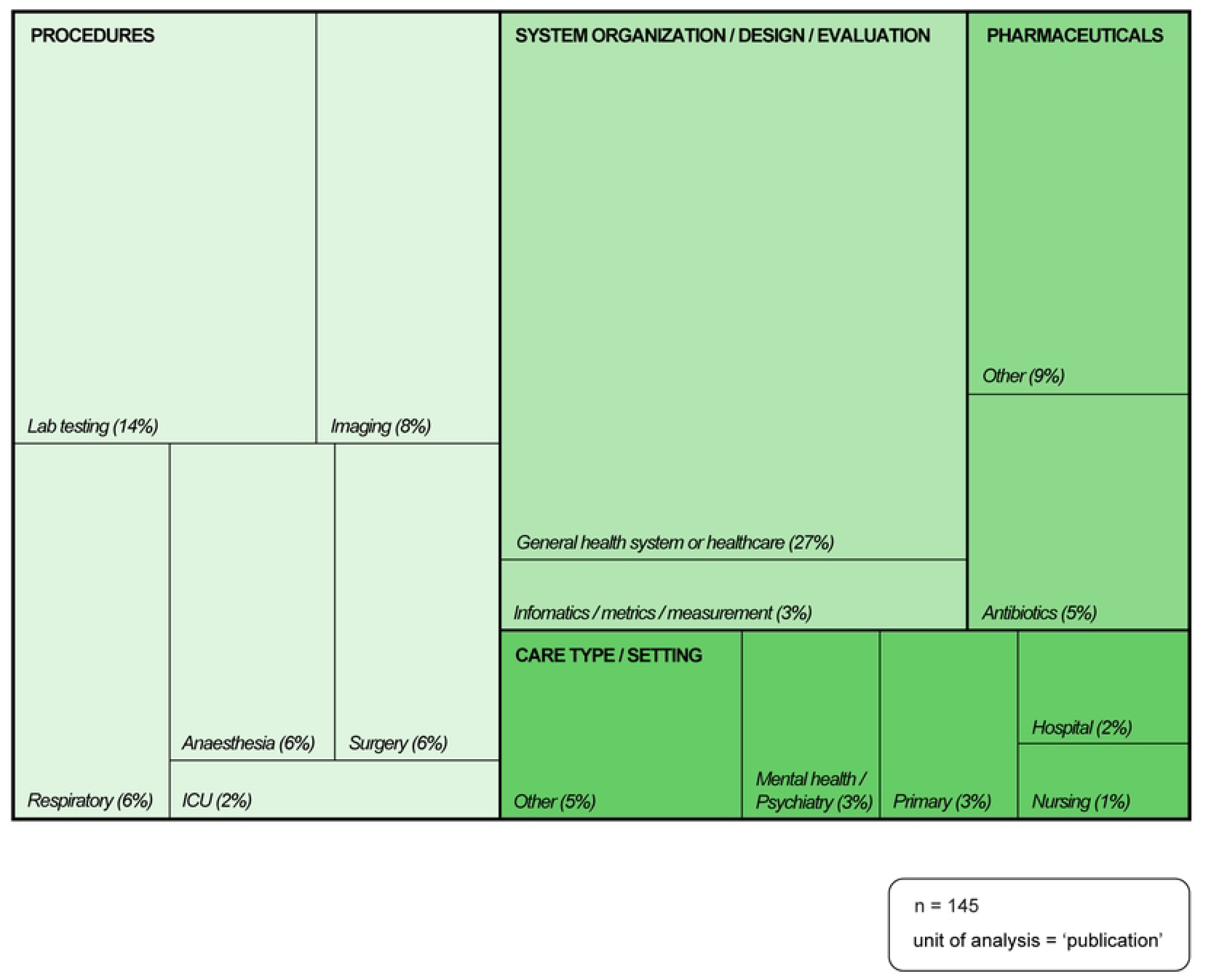
Healthcare focus of publications. Source: Microsoft Excel: analysisFig 5; Adobe Illustrator: visualization.

### Environmental Focus of Publications

As discussed in the methods section, we collected data related to environmental sustainability for the subset of empirical papers we categorized as (approximately) equally focused on environmental sustainability of healthcare and reducing LVC. For the 13 empirical studies included in this analysis, we reported results across six categories ‘*GHG emissions’*, *‘Pollution’, ‘Resource use’*, *‘Waste management*’, *‘Supply chain & facility/service design’* and *‘Environmental stewardship’* and 16 sub-categories of environmental outcome (Figure 4. below).

**Figure 4.**
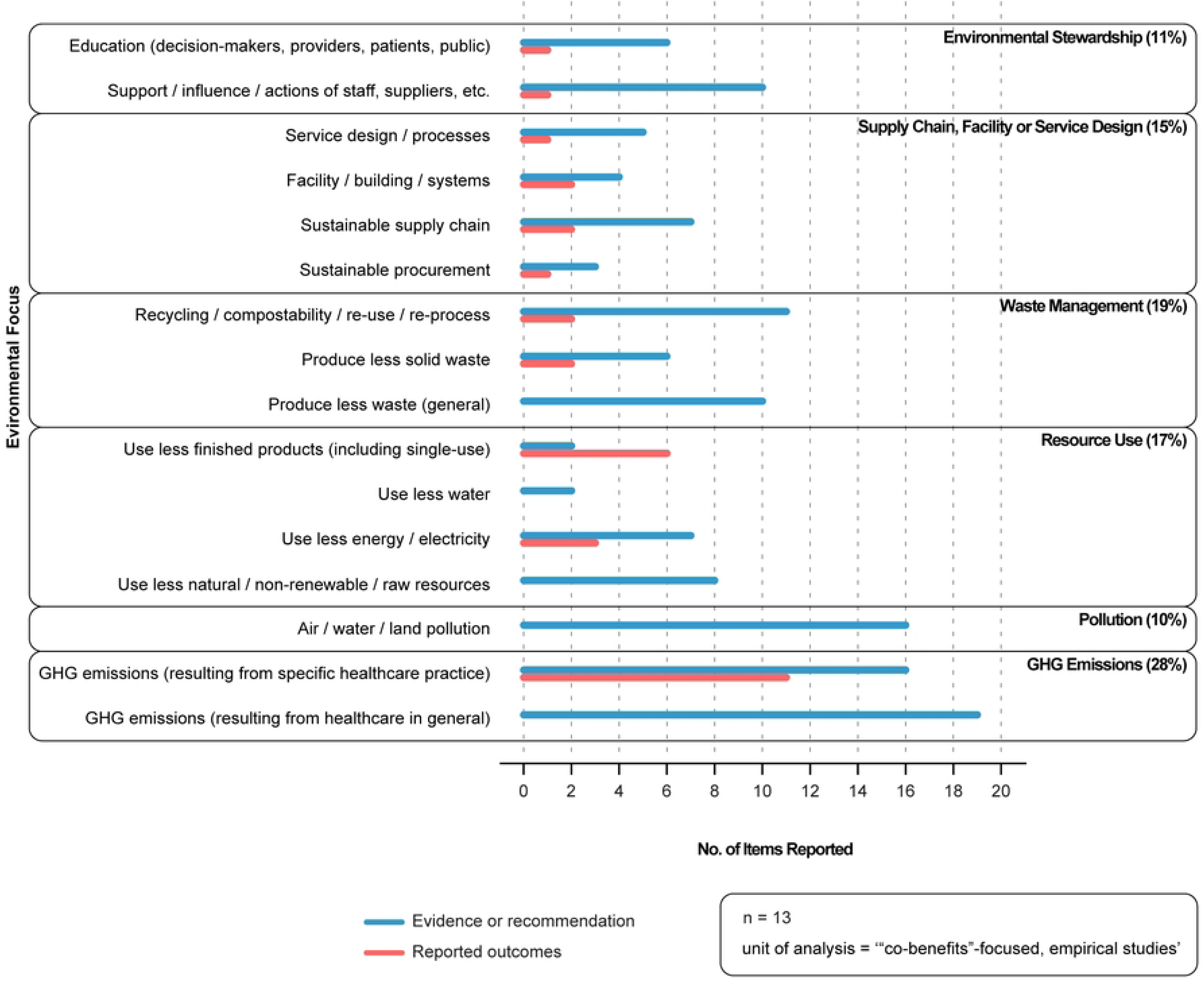
Environmental focus of publications. Source: Microsoft Excel: analysis; Adobe Illustrator: visualization.

As described in the methods section, within each of the sub-categories, data were classified as: (i) *evidence* or *recommendation*, if a study cited existing evidence or made a recommendation from an environmental sustainability category; or (ii) *reported outcomes*, if a study reported new study outcomes in an environmental sustainability category. Evidence or recommendation data were reported across all 16 sub-categories. The majority were for *‘GHG emissions’ (healthcare general)’, ‘GHG emissions (specific healthcare practice)’* and *‘Pollution’.* Reported outcomes were present in 11 of the 16 subcategories. Of the 13 studies that reported outcomes, seven reported outcomes across multiple categories; four reported a single outcome. Eleven of the 13 studies reported reduction in ‘*GHG emissions’*, followed by outcomes for ‘*Use less single-use products’*, then outcomes for *‘Use less energy’*.

## BIBLIOMETRIC ANALYSIS-SPECIFIC RESULTS

### Top Author Production Over Time

Five hundred eight-one unique authors contributed to the 145 included publications between 2013 and 2023 (July). There were 19 ‘top’ authors (authors with three or more publications), 14 of whom had already published relevant work in 2023. Most authors began publishing relevant publications from 2020 onwards. Seven of the ‘top’ authors were from the USA; six from Australia; three from Canada; two from the UK; and, one from the Netherlands.

### Author Collaboration Networks

Figure 5. depicts the top 10% of authors who have collaborated on at least one publication and describes the networked relationships between these authors. One node (grey coloured circle) represents one author; a solid grey line represents at least one collaboration between a pair of authors *within* a network (cluster of nodes); a dashed grey line represents at least one collaboration between a pair of authors *across* networks. The closer the nodes, the stronger the collaborative relationship. Nine distinct networks were identified. The largest networks, by number of collaborators involved (13 each), were centered around Forbes McGain in Australia (whose included publications were published between 2019 – 2023; and, who collaborated with all authors in the network) and Jodi Sherman in the USA (whose included publications were published between 2020 – 2023; and, who collaborated with all but one author in the network).

**Figure 5.**
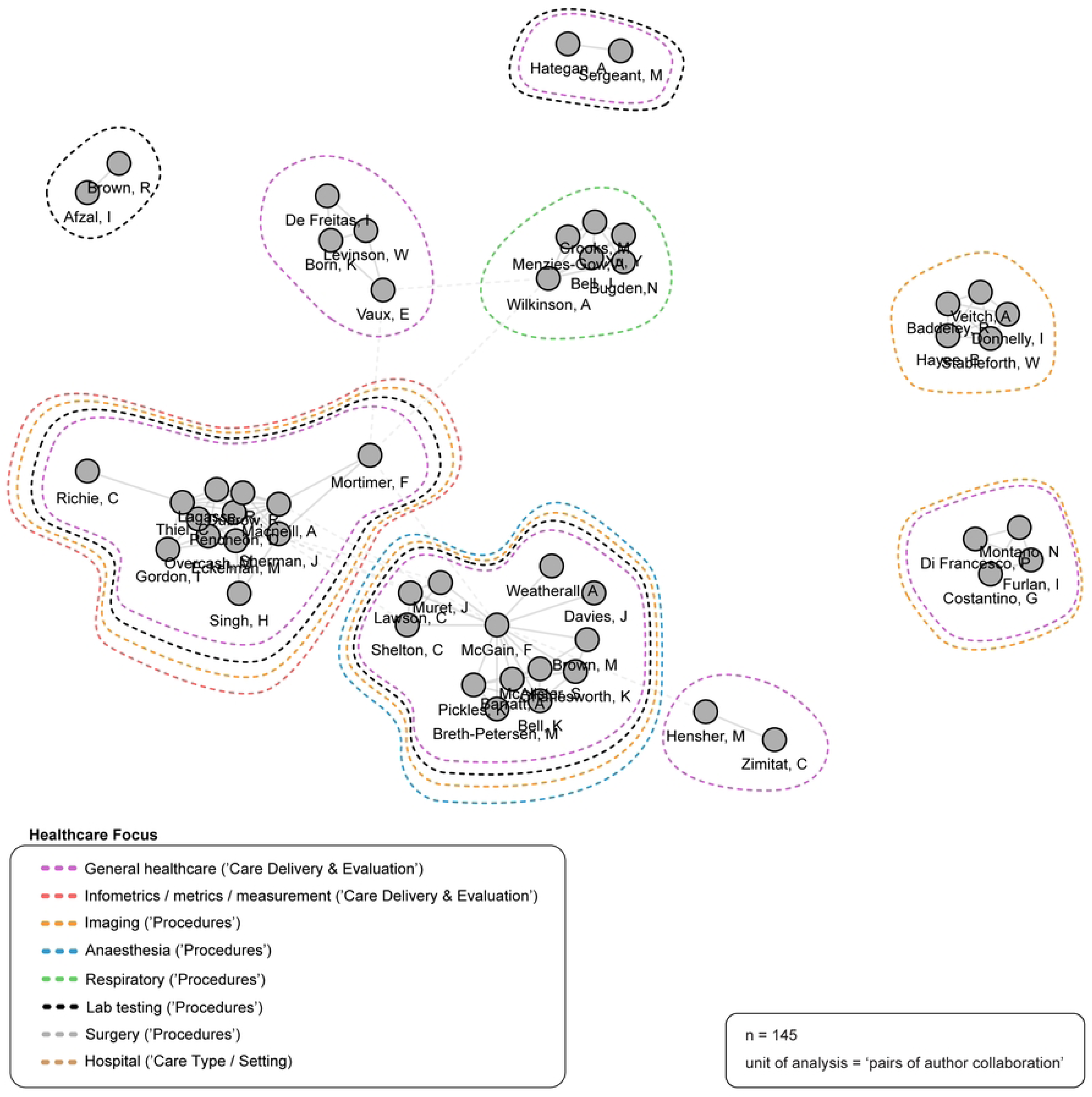
Author collaboration networks. Source: R-based application Biblioshiny was used; additional analysis (i.e., healthcare focus of publications) completed by the study authors and applied to the visualization using Adobe Illustrator. Parameters specified: analysis: collaboration network; field: authors; network layout: automatic (default); clustering algorithm: walktrap (default); normalization: association (default); number of nodes: 58 (top 10% of total authors); repulsion force: 0.1 (default); remove isolated nodes: yes (default); minimum number of edges: 1 (default).

### Country Collaboration Networks

Figure 6. depicts the country collaboration networks based on author institutions. One node (coloured circle) represents one country. The closer the nodes, the stronger the collaborative relationship. Two primary networks were identified (blue, red). The largest network by number of countries captured (blue) was comprised of 17 unique countries, and represents seven publications published between 2019 – 2023 and generated by two or more of the authors within the network (but not exclusively). The second largest, but most productive, network (red) was comprised of 11 unique countries, and represents 24 publications published between 2019 – 2023 and generated by two or more of the authors within the network (but not exclusively). The remaining networks (purple, green, orange, pink, brown, grey) each represent one publication (n = 6) published between 2019 – 2023 and generated through collaboration with authors situated within the primary networks (blue, red).

**Figure 6.**
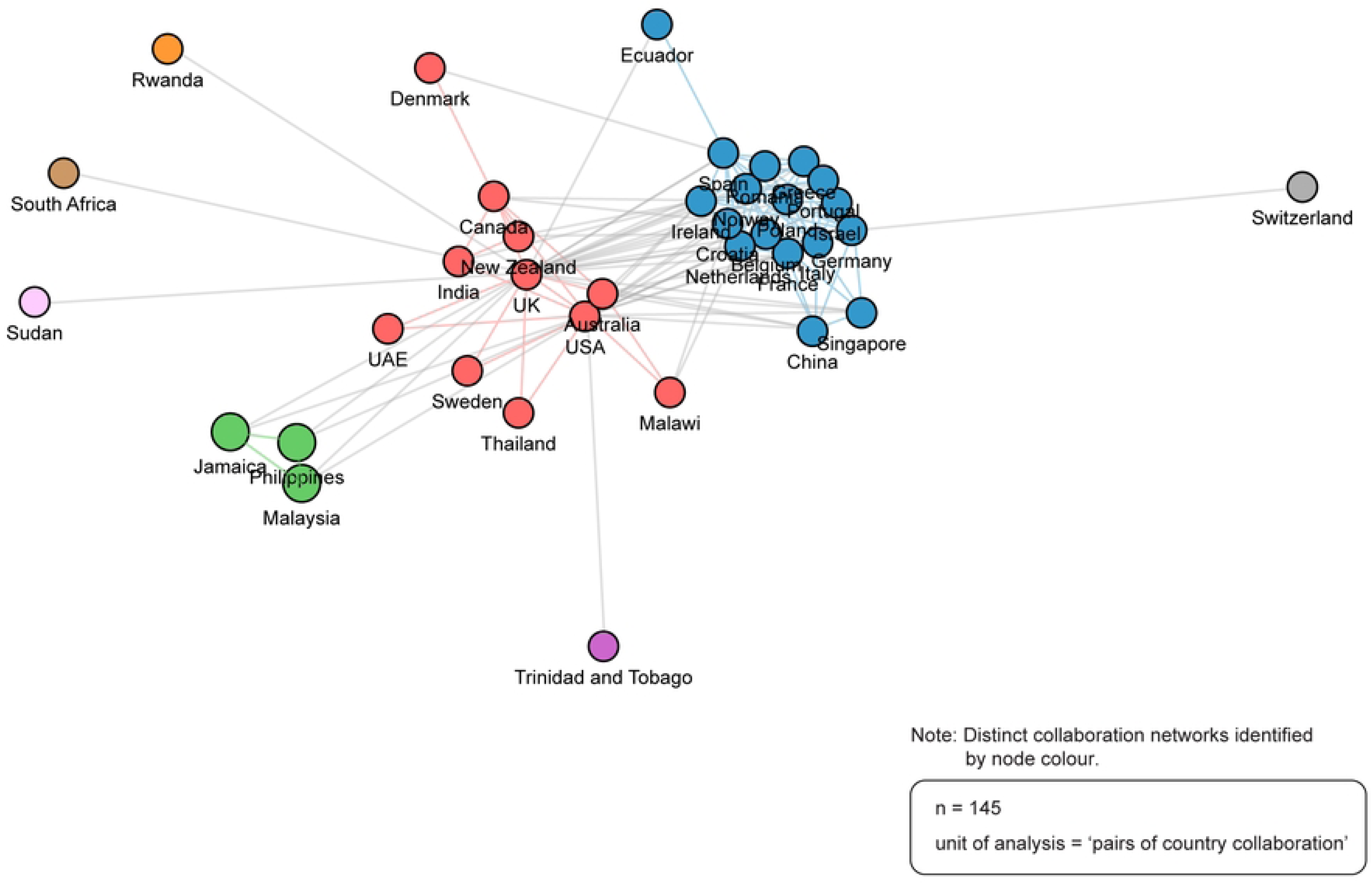
Country collaboration networks. Source: R-based application Biblioshiny was used; additional analysis (i.e., number of publications generated by each network) completed by the study authors; visualization recreated using Adobe Illustrator. visualization recreated using Adobe Illustrator. Parameters specified: analysis: collaboration network; field: countries; network layout: automatic (default); clustering algorithm: walktrap (default); normalization: association (default); number of nodes: 200 (all countries in world, rounded); repulsion force: 0.1 (default); remove isolated nodes: yes (default); minimum number of edges: 1 (default).

### Top Institutions

Three hundred eighty-seven unique institutions (affiliates) contributed to the 145 included publications. Figure 7. depicts the top institutions (with four or more publications). There were 16 ‘top’ institutions: five were from Australia; four from the USA; four from the UK; and, three from Canada. The top-producing institution was the University of Sydney with 18 publications. The majority of top-producing institutions were universities, followed by medical centres or medical organizations.

**Figure 7.**
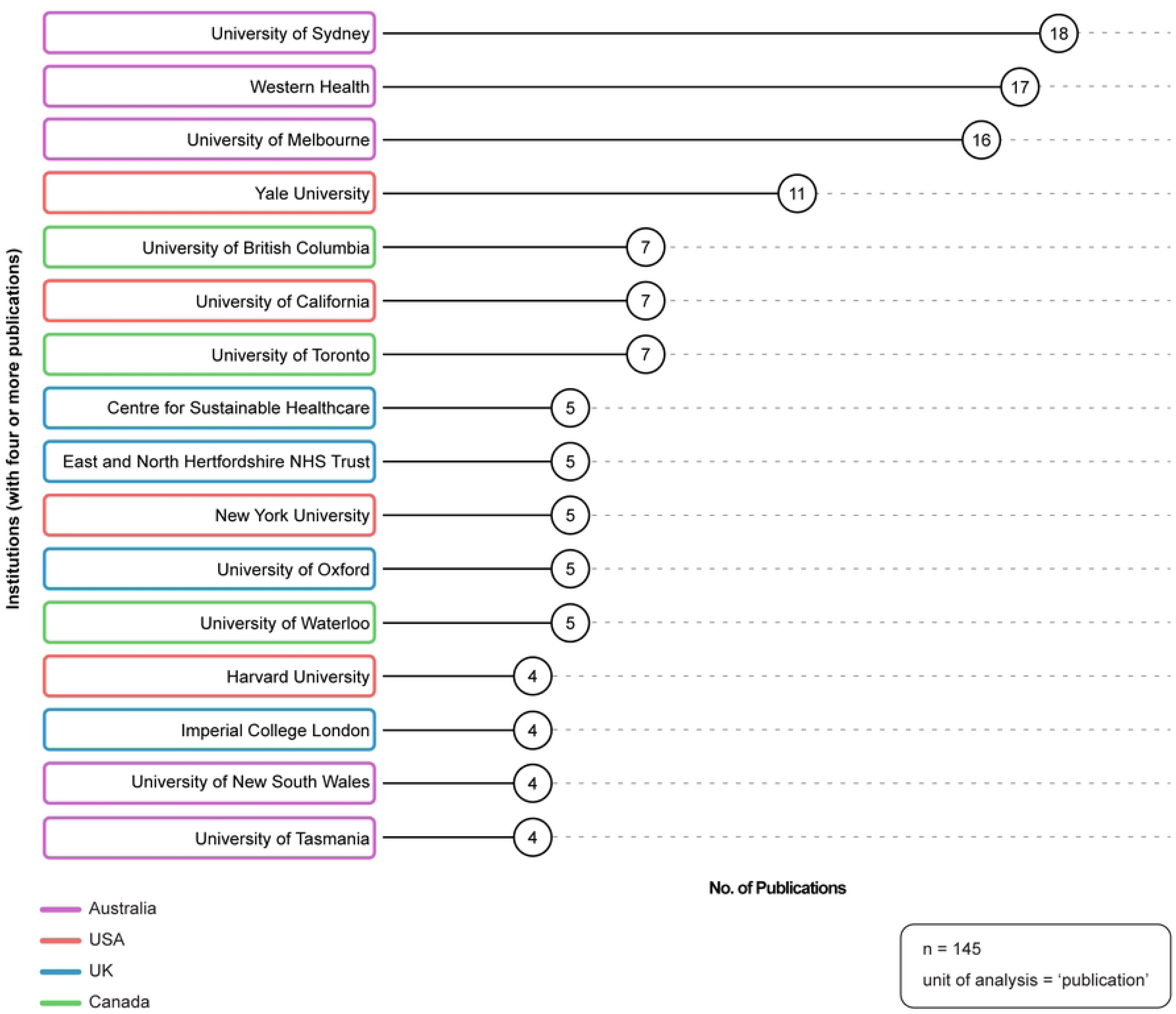
Top institutions. Source: R-based application Biblioshiny was used; additional analysis (i.e., institution country of origin; institution-type) completed by the study authors and applied as necessary to the visualization using Adobe Illustrator. Parameters specified: analysis: most relevant affiliations; affiliation name disambiguation: no; number of affiliations: 16.

### Top Journals

One hundred and seven unique journals contributed to the 145 included publications. There were six ‘top’ journals (journals with three or more publications). ‘BMJ’ was the top-producing journal, with 12 publications; followed by ‘Healthcare Papers’ and ‘Journal of Climate Change and Health’ with four each; ‘Medical Journal of Australia’, ‘Resources Conservation and Recycling’ and ‘Social Science and Medicine’ with three each.

## DISCUSSION

The results of this study provide important insights into the emerging literature on the environmental co-benefits of reducing LVC. This section offers a detailed discussion of the key findings, applications, and directions for future work.

The publication trend over time demonstrated that research and evaluation to inform practice changes in this area has dramatically increased over the last three years. The fact that there were more publications in first half of 2023 than in all of 2022, and that the majority of the top authors have published in the first half of 2023, shows significant momentum within the field. While the majority of publications to date are commentaries/perspectives/opinions; this characteristic is typical of an emerging field. The field will benefit from more empirical studies to assess the environmental harms resulting from healthcare and how these harms are reduced or eliminated when LVC is reduced or eliminated. Future empirical work should endeavour to conduct rigorous investigations and measure and report on the broad spectrum of environmental harms.

The collaboration network analyses revealed that large, international groups of authors are working together to advance this field. The largest author collaboration networks are centered around Australian and American authors and the country collaboration network analysis revealed that the most productive networks were spearheaded by authors in the UK, USA, Canada and Australia. In addition, the results show that in recent years, increasingly, a greater number of authors from countries with emerging economies (such as Thailand, India, Jamaica, and Sudan) have been contributing to knowledge production in this field. While the community doing work in this field is small and quite concentrated in a few countries and institutions, the volume and momentum behind this work provides a significant opportunity for knowledge sharing and consensus development.

Our findings also demonstrated that the majority of included publications focused on environmental sustainability while flagging the importance of reducing LVC as a possible strategy. Importantly, the number of publications categorized as ‘(approximately) equally’ focused on the environmental co-benefits of reducing LVC has increased steadily over time and these publications arguably best advance the agenda of realizing the environmental co-benefits of reducing LVC. Scholarship is beginning to present the importance of reducing LVC and environmental harms as inextricably linked. Thiel & Ritchie [12] describe the pernicious cycle of “harm, treat, harm” to describe the paradox of healthcare harming the environment, which in turn harms human health and requires more healthcare, which further harms the environment. For example, air pollution is known to induce breathing difficulties, and inhalers are used to minimize the effects of air pollution, but inhaler-use generates a significant amount of carbon dioxide, which then exacerbates air pollution [12]. This cycle makes reducing LVC even more poignant, as care that produces environmental harms—but no benefits to patients—represents the antithesis of healthcare’s mandate.

The healthcare focus results highlighted that the included publications covered a broad scope and diverse practices in healthcare. To capture this diversity, we developed 15 ‘healthcare focus’ sub-categories as a framework to begin to organize this work across system organization/ design/evaluation, procedures, pharmaceuticals and care type/settings. While the scope was broad, the majority of publications were categorized as focused on general healthcare. The included empirical publications focused on targeted practice change interventions for specific healthcare practices, primarily, reducing unnecessary laboratory testing and appropriate inhaler use. These two areas are also significant foci for research strictly focused on reducing environmental harms or reducing LVC and represent a logical merging for these fields.

The analysis of environmental sustainability focus demonstrated that the included empirical studies cited evidence, made recommendations, and reported outcomes across a broad spectrum of environmental sustainability outcomes. While we recognize the sample size was small and the categorization framework broad, our goal was to map essential aspects of environmental harm and efforts, through reducing LVC, to reduce it. The strong focus on GHG emissions in the empirical studies is likely due to the fact that GHG emissions are the most available environmental data in healthcare and can be translated into relatable results (e.g., equivalency to driving distances). While valuable, this focus also highlights the need to develop ways to evaluate all environmental sustainability impacts, collect rigorous data, monitor and report on broader outcomes. In addition to GHG emissions, studies focused on solid waste management and recycling which are established areas of environmental improvement. Of note, the included studies also extended into important, emerging areas of sustainability such as composability, reprocessing or reuse [19, 32, 33]. Sustainable supply chain and procurement [32, 34] and environmental stewardship [32, 33, 35] are also critical environmental harms beginning to receive attention in this literature. We acknowledge that the outcomes we have categorized (using less resources, better waste management, more sustainable procurement) will all result in less GHG emissions, but it is important to explicate these processes as healthcare providers and organizations need to develop specific interventions to address these environmental harms.

Reducing LVC is a critical strategy to bolster health system sustainability and the recognition of the environmental co-benefits of reducing LVC is gaining prominence and momentum. Thiel and Richie [12] describe ‘rejecting health care overuse’ as an act of beneficence by reducing both patients and global citizens’ risk of climate change induced health harms. As mentioned, the majority of LVC occurs in HIC settings, while environmental harms disproportionally impact LMIC settings [20, 21]. It is also important to note emerging research that identifies increasing overuse issues in low-resource settings [36]. The pervasiveness of LVC further supports global research and evaluation to inform practice change in this area, as overuse in LMIC settings is further exacerbated by limited public budgets, access to resources and complex population health needs [36].

## LIMITATIONS

This study has several limitations. The search strategy was complicated by numerous factors such as no agreed upon search terms or MeSH terms, and the inability to search on terms like ‘environment’, ‘sustainable’, waste’, inappropriate’ (as these terms have broad meaning in healthcare and pull millions of results). To mitigate these issues, we iteratively developed the search strategy. While we acknowledge that our final set of ‘environmental’ terms appears to be very carbon-focused, we feel confident that our search was exhaustive. Publications in languages other than English were not included due to a lack of resources. Finally, while there is no clear consensus in the bibliometric analysis guidance literature regarding the generalizability of bibliometric study outputs to the field under study, we believe the systematic, comprehensive, and detailed approach used to identify relevant publications provides a robust understanding of the “co-benefits” field, despite not necessarily representing a “complete” population.

## CONCLUSION

This foundational study is an important first step to identify research and evaluation to inform practice change on the environmental co-benefits of reducing LVC and authors and institutions doing this important work. By systematically and comprehensively collecting and analysing data on this emerging field, our research supports evidence-based health system improvement work with the potential to increase effectiveness and efficiencies in resource constrained health systems. Future research should focus on conducting rigorous empirical studies in this area, including evaluation and reporting on the broad spectrum of environmental harms.

## Data Availability

All relevant data are within the manuscript and its Supporting Information files.

## SUPPLEMENTARY MATERIAL

Supplementary File 1. PRISMA-ScR Checklist

Supplementary File 2. Preferred Reporting Items for Systematic Bibliometric Analysis (PRISBA) Checklist

Supplementary File 3. Medline Search Strategy

Supplementary File 4. Study methods overview

Supplementary File 5. Bibliometric analysis data cleaning actions Supplementary File 6. Included publications

Supplementary File 7. Bibliometric analysis dataset

## ACKNOWLEDGEMENTS

The authors would like to thank Tasneem Bohra for her valuable assistance.

## Author Contributions

GP and FAM conceived of the study. GP, KB, and FAM developed the study protocol. GP and SH identified relevant publications. GP and SH extracted and analyzed the data and generated the data visualizations; KB and FAM provided critical feedback throughout these processes. GP and SH drafted the manuscript. GP, SH, KB, and FAM revised the draft manuscript for publication.

## Disclaimer

No funders provided input into the content of this manuscript.

## Patient and public involvement

Patients and/or the public were not involved in the design, or conduct, or reporting, or dissemination plans of this research.

